# Cross-ancestry genetic investigation of schizophrenia, cannabis use disorder, and tobacco smoking

**DOI:** 10.1101/2024.01.17.24301430

**Authors:** Emma C Johnson, Isabelle Austin-Zimmerman, Hayley HA Thorpe, Daniel F Levey, David AA Baranger, Sarah MC Colbert, Ditte Demontis, Jibran Y Khokhar, Lea K Davis, Howard J Edenberg, Marta Di Forti, Sandra Sanchez-Roige, Joel Gelernter, Arpana Agrawal

## Abstract

Individuals with schizophrenia frequently experience co-occurring substance use, including tobacco smoking and heavy cannabis use, and substance use disorders. There is interest in understanding the extent to which these relationships are causal, and to what extent shared genetic factors play a role. We explored the relationships between schizophrenia (Scz), cannabis use disorder (CanUD), and ever-regular tobacco smoking (Smk) using the largest available genome-wide studies of these phenotypes in individuals of African and European ancestries. All three phenotypes were positively genetically correlated (r_g_s = 0.17 – 0.62).Causal inference analyses suggested the presence of horizontal pleiotropy, but evidence for bidirectional causal relationships was also found between all three phenotypes even after correcting for horizontal pleiotropy. We identified 439 pleiotropic loci in the European ancestry data, 150 of which were novel (i.e., not genome-wide significant in the original studies). Of these pleiotropic loci, 202 had lead variants which showed convergent effects (i.e., same direction of effect) on Scz, CanUD, and Smk. Genetic variants convergent across all three phenotypes showed strong genetic correlations with risk-taking, executive function, and several mental health conditions. Our results suggest that both horizontal pleiotropy and causal mechanisms may play a role in the relationship between CanUD, Smk, and Scz, but longitudinal, prospective studies are needed to confirm a causal relationship.

## Introduction

Schizophrenia (Scz) is a psychiatric condition with an estimated twin-based heritability of around 80%^1,2^. Substance use disorders (SUDs) are highly prevalent in individuals with Scz^3^. Of these co-occurring SUDs, the role of cannabis use as a risk factor for Scz and first episode psychosis onset remains a classical “chicken or egg” problem in psychiatry^4^.

Some studies have suggested a causal, dose- and age-dependent effect of cannabis use on risk for onset of Scz and other forms of psychosis^5–7^. However, cannabis use and cannabis use disorder (CanUD) are heritable^8^ (twin heritability ∼50%), and an alternative hypothesis is that shared genetic pathways underlie liability to Scz and cannabis use phenotypes^9,10^. Genetic correlations from genome-wide association studies (GWAS) have provided support for some genetic commonality (e.g., SNP-r_g_ (Scz, cannabis use) = 0.25^11^, SNP-r_g_ (Scz, CanUD) = 0.37^12^). A recent study identified 27 and 21 genome-wide significant loci contributing to the shared genetic etiology between Scz and cannabis use and CanUD, respectively^13^. However, the identification of shared loci was largely driven by genome-wide significant loci in the Scz GWAS, due to the relative difference in discovery power between the Scz and cannabis GWASs. Furthermore, these prior studies have largely been performed in samples predominantly of European ancestry, limiting the generalizability of these findings.

Horizontal pleiotropy (i.e., genetic variants independently contributing to both CanUD *and* Scz) *and* vertical pleiotropy (i.e., shared genetic associations via a causal path) are not mutually exclusive; both mechanisms may play a role in the co-occurrence of CanUD and Scz. Genetically informed studies of CanUD and Scz have reached mixed conclusions, with no single direction of causality receiving overwhelming support^14,15^. Several Mendelian Randomization analyses have suggested greater support for Scz causing cannabis use and CanUD than the opposite direction^11,16^, while the most recent GWAS of CanUD found a bidirectional causal association between Scz and CanUD^17^.

Few prior genetic studies have attempted to disentangle how nicotine/tobacco use genetics impacts the genetic relationship between Scz and CanUD. Approximately 72% of those with Scz report daily tobacco smoking (while this same report estimated 43% were regular cannabis users^18^), and there is evidence that individuals who smoke tobacco daily are at increased risk of psychosis^19^, an earlier age of onset of first psychotic episode^19^, and the development of schizophrenia^20^. The prevalence of tobacco use, whether as tobacco cigarettes or consumed with cannabis in certain preparations (e.g., blunts, where tobacco is removed from a cigar and replaced with cannabis, or spliffs, where cannabis and tobacco are rolled together), is also high in individuals with CanUD^21,22^. Prior studies have reported genetic correlations of tobacco smoking with CanUD (SNP-r_g_ = 0.61^17^) and Scz (SNP-r_g_ = 0.14^23^). Despite this, few epidemiologic studies have taken potential genetic sharing into account when reporting evidence for causal relationships between tobacco, cannabis, and Scz^6,7^. In turn, few genomic studies of cannabis and Scz have considered the role of tobacco^13^, despite the frequent co-occurrence of tobacco and cannabis use, especially in Europe^24^. In a prior study, we found that genetic liability for CanUD was positively associated with genetic liability for Scz even when accounting for the genetic components of cannabis ever-use, tobacco smoking, and nicotine dependence^10^. Another study found a causal effect of genetic liability to cannabis use on risk for schizophrenia, and this association was unchanged when accounting for tobacco smoking^15^. Thus, the genetic association between cannabis and Scz appears to be independent of tobacco use genetics to some extent, although the relatively low power of prior CanUD GWAS meant limited conclusions could be drawn from these earlier studies.

Given the significant genetic correlations between CanUD, tobacco smoking, and Scz, the increasing pace of cannabis legalization with emerging increases in CanUD incidence^25^, parallel increases in the popularity of nicotine vaping^26^, and the consequent potential impact on the course of Scz in those with heavy cannabis and tobacco use^27–31^, we investigated the evidence for causal relationships and horizontal pleiotropy between CanUD, tobacco smoking, and Scz. We used the largest genome-wide summary statistics available for Scz^32^ (European ancestry N = 161,405; African ancestry N = 15,846), CanUD^17^ (European ancestry N = 886,025; African ancestry N = 120,208), and ever-regularly smoking tobacco^33^ (Smk; European ancestry N = 805,431; African ancestry N = 24,278) in samples whose genetic ancestry is most similar to those historically from Europe (henceforth referred to as “European ancestry”) and samples whose genetic ancestry is most similar to those historically from Africa (henceforth referred to as “African ancestry”) to identify and characterize pleiotropic signals and conduct causal inference analyses. We focused on CanUD and Smk (as opposed to cannabis ever-use, or cigarettes per day) as CanUD was the cannabis phenotype with the largest genetic correlation with Scz, there was no available GWAS of cannabis consumption or heaviness of use, and current GWAS of nicotine dependence (relying on the Fagerström Test for Nicotine Dependence^34^ (FTND)) have been relatively under-powered compared to Smk.

## Results

### Genome-wide genetic correlations

Schizophrenia (Scz), Cannabis Use Disorder (CanUD), and ever-smoking tobacco regularly (Smk) were significantly genetically correlated in the European ancestry data (**Table S1**). The magnitude of the genetic correlation between Scz and CanUD (r_g_ = 0.37, SE = 0.02, p= 2.97e-60) was statistically greater (p_diff_ = 6.5e-18) than the correlation between Scz and Smk (r_g_ = 0.17, SE = 0.02, p = 6.88e-20) or between Scz and a measure of nicotine dependence more similar to CanUD, the FTND (r_g_ = 0.22, SE = 0.04, p = 1.56e-7; p_diff_ = 0.002). This suggests that our choice of ever-regular smoking, rather than the FTND, as a measure of tobacco use was not the reason for the lower genetic correlation.

In the African ancestry data, the largest genetic correlation was between Scz and CanUD (r_g_ = 0.61, SE = 0.14, p = 1.41e-5; **Table S1**). While the genetic correlation between Scz and Smk (r_g_ = 0.34, SE = 0.15, p = 0.03) was of greater magnitude than in the European ancestry data, this estimate was not significantly different from zero after accounting for multiple testing, due to the much larger standard error.

### Causal inference analyses

Using CAUSE^35^, a method that accounts for both correlated and uncorrelated horizontal pleiotropic effects, we found evidence for bidirectional causal relationships between all three phenotypes in the European ancestry data (**Figure 1a, Table S2**).

**Figure 1.**
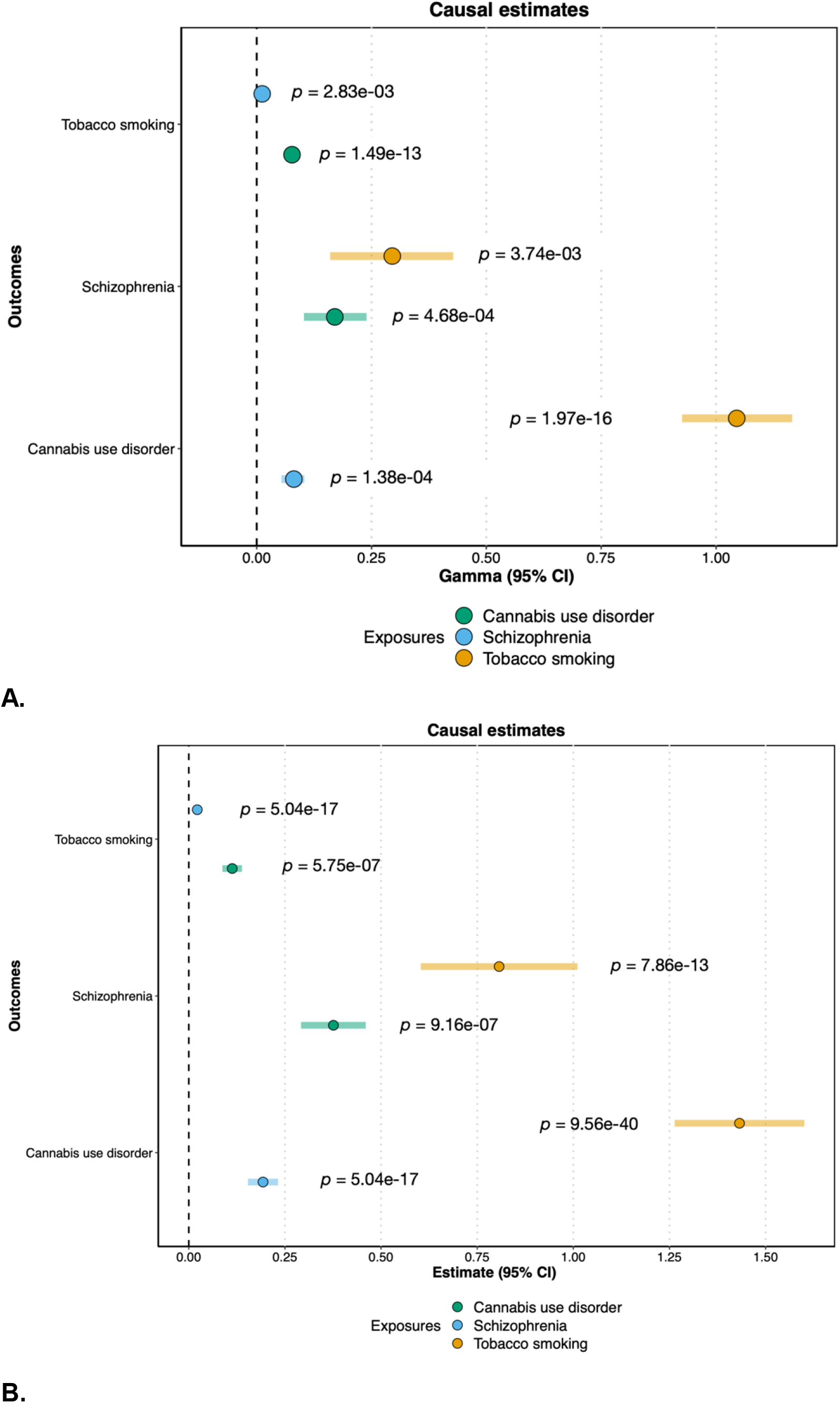
Panel A: Causal estimates (gamma) and 95% confidence intervals from CAUSE. Panel B: Causal estimates (beta) and 95% confidence intervals from MR-PRESSO. “Exposure” phenotypes are indicated by the color, while “Outcome” phenotypes are listed on the y-axis.

To explicitly test for the presence of horizontal pleiotropy, and to ensure our results were not isolated to a specific method of causal inference, we also performed Mendelian Randomization Pleiotropy RESidual Sum and Outlier (MR-PRESSO^36^) analyses in the European ancestry data. The MR-PRESSO global test for horizontal pleiotropy was significant for each pairwise test, and we found significant bidirectional causal effects between all three traits after the removal of outliers for horizontal pleiotropy (**Figure 1b**), consistent with the results from CAUSE. Results from other MR methods were generally consistent, with the same direction of effect (**Table S3**), although the more conservative MR-Egger test^37,38^ only showed a statistically significant causal effect of Smk on CanUD.

### Cross-trait loci: European ancestry

In consideration of the significant genetic correlations and evidence for horizontal pleiotropy from MR-PRESSO, we used ‘Association analysis based on SubSETs’ (ASSET^39^) to combine the GWAS summary data for CanUD, Smk and Scz (separately by ancestry), using the two-tailed meta-analysis approach. Unlike traditional meta-analysis approaches, ASSET accounts for SNPs with significant effects on multiple disorders even if the effects on the traits are in opposite directions. Following Lam et al.^40^, we use the following notation for each subset: ∩ represents variant subsets with the same directions of effect (+ or -), and | represents variant subsets whose effects are divergent across the different phenotypes. We therefore defined four subsets: (1) Scz ∩ CanUD ∩ Smk (i.e., a subset with convergent effects across all 3 traits); (2) Scz ∩ CanUD | Smk (i.e., a subset of variants with convergent effects for Scz and CanUD, but divergent effects for Smk); (3) Scz ∩ Smk | CanUD; and (4) CanUD ∩ Smk | Scz.

In total, we identified 439 pleiotropic genomic risk loci (i.e., loci where the lead SNP has an effect on all three phenotypes). Of these, 150 loci were novel (i.e., not genome-wide significant in any of the original GWAS; see **Table S4** and **Table S5**), with 127 of these loci having lead SNP p-values ≤ 1e-5 in at least one of the original GWAS, and the remaining 23 having p-values ≤ 1.4e-4.

For the subset of SNPs with convergent effects across all 3 traits (Scz ∩ CanUD ∩ Smk) in the European ancestry samples, we identified 202 genomic risk loci with 259 lead SNPs (**Table S6**). The strongest locus was on chromosome 8, with the top lead SNP being rs73229090 (chr8:27442127, p = 1.5e-62; **Figure 2**), located in an intron of the non-coding gene *GULOP*, replicating previous associations with each trait (e.g., ^41–43^). This SNP is also an expression quantitative trait locus (eQTL) for *EPHX2* in B cells, tibial artery, esophagus, and cultured fibroblast cells, *CHRNA2* in the cerebellum, and *CCDC25* in the nucleus accumbens.

**Figure 2.**
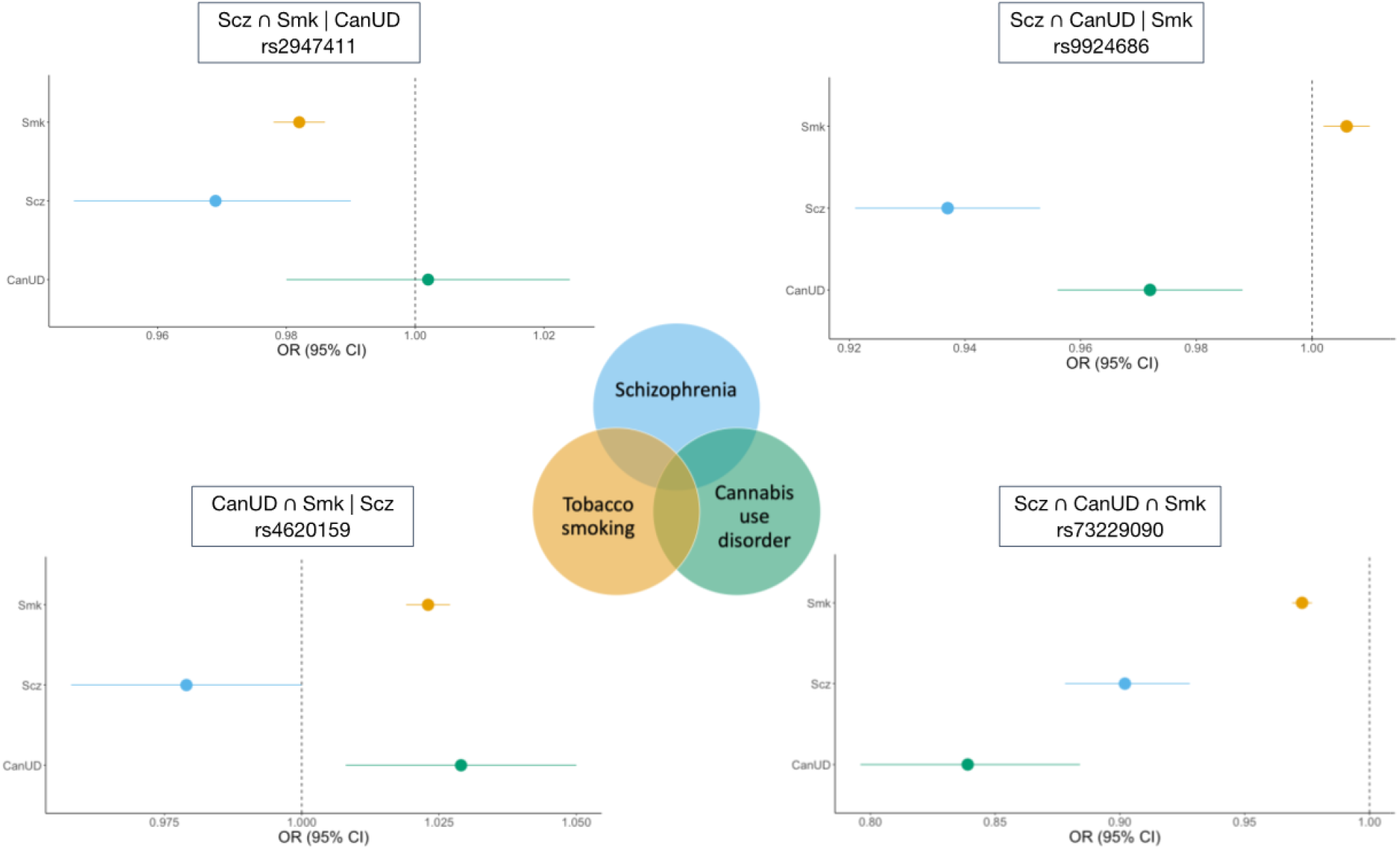
Example forest plots from the ASSET European ancestry cross-disorder meta-analysis of CanUD, Smk, and Scz. The lower right panel shows lead SNP (rs73229090) in Scz ∩ CanUD ∩ Smk subset. The upper right panel shows SNP (rs9924686) in Scz ∩ CanUD | Smk subset. The upper left panel shows SNP (rs2947411) in Scz ∩ Smk | CanUD subset. The lower left panel shows SNP (rs4620159) in CanUD ∩ Smk | Scz subset.

The Scz ∩ CanUD | Smk subset of SNPs revealed 37 genomic risk loci with 37 lead SNPs (**Table S7**). The top association was on chromosome 16, with lead SNP rs9924686 (chr16:30003076, p = 3.3e-15) within a locus previously implicated by Scz GWAS^41^. This SNP, located in the 3’ untranslated region of the serine/threonine-protein kinase gene *TAOK2*, has a CADD score of 18.16, suggesting deleteriousness, and a RegulomeDB score of 1f (eQTL + transcription factor binding/DNase peak), suggesting that this SNP is likely to affect transcription factor binding and linked to expression of a gene target. Furthermore, rs9924686 is an eQTL for several genes, including genes associated with metabolic and immunological traits^44,45^ (*YPEL3* and *INO80E* in adipose tissue and several brain tissues) and alcohol intake^45,46^ (*PPP4C* and *MVP* in cultured cell fibroblasts).

We identified 46 genomic risk loci with 48 lead SNPs for the Scz ∩ Smk | CanUD subset (**Table S8**). Chromosome 2 had the strongest signal in this subset, with intergenic lead SNP rs2947411 (chr2:614168, p = 3.6e-19) that replicates previous associations with Smk^43^. This SNP was an eQTL for only one gene (*SH3YL1* in whole blood).

There were 114 genomic risk loci and 143 lead SNPs for the CanUD ∩ Smk | Scz subset (**Table S9**). The strongest meta-analytic effect was at lead SNP rs4620159 on chromosome 6 (chr6:111744735, p = 1.8e-28); this locus was previously associated with Smk and CanUD^47,48^. The lead SNP is an intronic variant in *REV3L*, a gene previously associated with smoking and several metabolic traits^23,45^.

### Cross-trait loci: African ancestry

No associations passed the genome-wide significance threshold (alpha = 5e-8) in the ASSET analysis of the African ancestry data. However, the 14,001 pleiotropic SNPs that were genome-wide significant in the European ancestry data showed smaller p-values than expected by chance in the African ancestry data (i.e., the distribution of p-values was significantly left-skewed, with a Kolmogorov-Smirnov goodness-of-fit test indicating significant (p ≤ 2e-16) divergence from a distribution of 14,001 randomly sampled SNP p-values). This suggests that with larger sample sizes, future analyses might identify similar loci across both the European and African ancestry datasets.

### Cross-ancestry meta-analysis

We performed a sample size-weighted cross-ancestry meta-analysis of the ancestry-specific *one-sided* meta-analysis results from ASSET. Unlike the ancestry-specific two-tailed meta-analyses described above, the one-sided meta-analysis in ASSET is more like a traditional meta-analysis, resulting in one effect size per SNP regardless of whether the SNP shows divergent directions of effect across traits. The cross-ancestry meta-analysis of CanUD, Smk, and Scz resulted in 448 genome-wide significant risk loci (**Table S10**).

### Genetic associations with other phenotypes

After defining SNP subsets using ASSET, we used GNOVA^49^ to estimate genetic correlations between the SNP subsets and educational attainment^50^ (Edu), executive function^51^, risk-taking^46^, and Townsend deprivation index (TDI; a regional measure of deprivation in the UK) in the European ancestry data (**Figure 3, Table S11**). Edu has previously been shown to be positively correlated with a subset of variants contributing to Scz risk^40^, despite negative genetic correlations between Scz and cognitive function^52^, and we expected that related socioeconomic status (i.e., TDI), executive function, and risk-taking phenotypes might be differentially associated with SNP subsets. For all subsets, the effect estimate was aligned with the direction of effect for CanUD.

**Figure 3.**
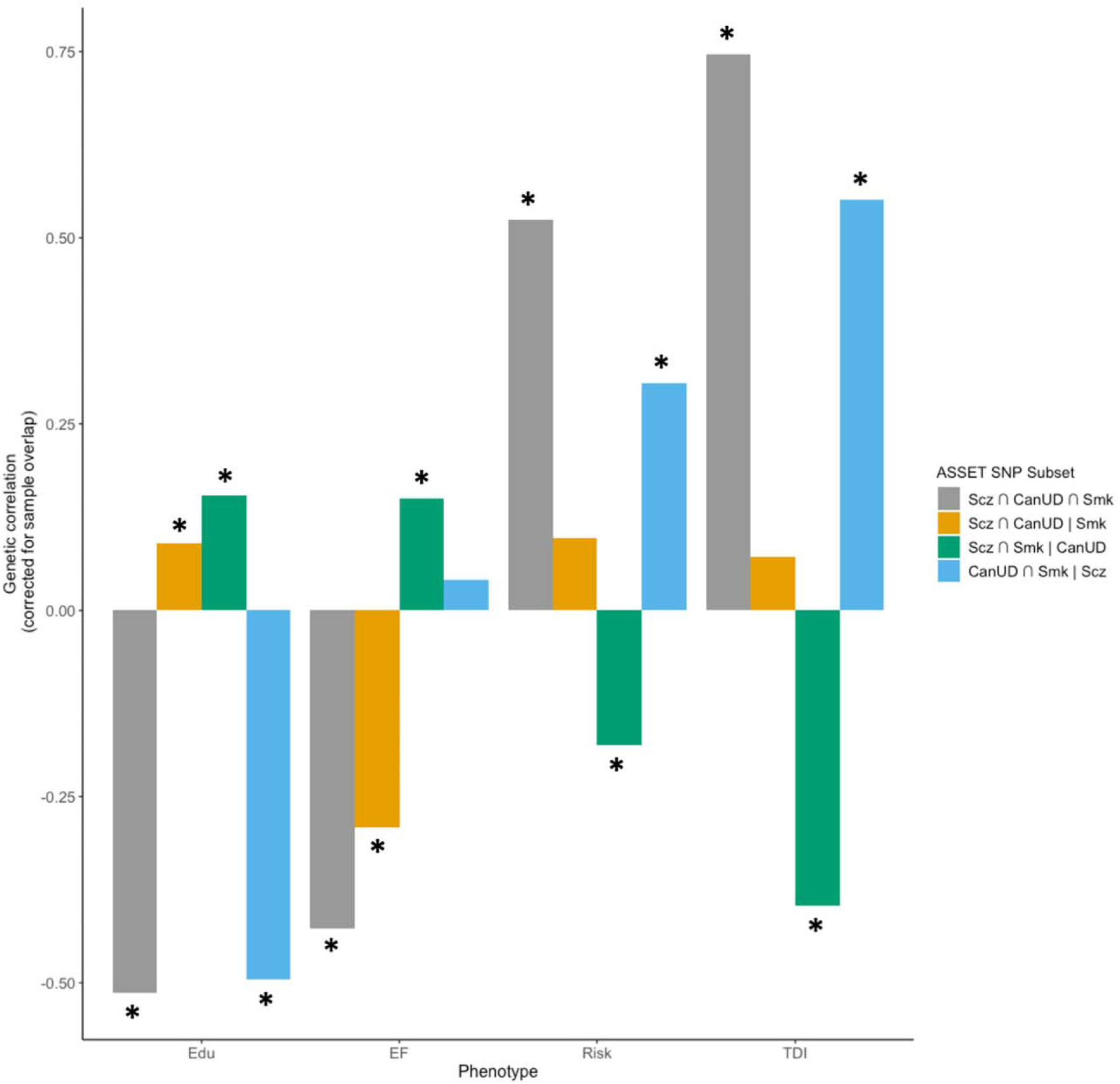
Estimated genetic correlations between educational attainment (Edu), executive function (EF), risk-taking (Risk), and Townsend deprivation index (TDI) and SNP subsets from ASSET. Asterisks (^*^) represent genetic correlations that are statistically significant after Bonferroni correction for 16 tests (p < 0.003).

The Scz ∩ CanUD ∩ Smk subset (i.e., variants with the same direction of effect on all three phenotypes) showed the strongest genetic correlations with all traits tested except Edu, where the Scz ∩ CanUD ∩ Smk and CanUD ∩ Smk | Scz subsets showed similar magnitudes of genetic correlation. For Edu, risk-taking, and TDI, the Scz ∩ CanUD ∩ Smk and CanUD ∩ Smk | Scz subsets showed the same direction of genetic correlation, while the Scz ∩ Smk | CanUD subset showed correlations in the opposite direction. In other words, genetic variants with the same direction of effect on CanUD and Smk, regardless of the direction of effect on Scz, showed similar negative genetic correlations with Edu, and positive genetic correlations with risk-taking and TDI, while genetic variants with the same direction of effect on Scz and Smk but not CanUD showed correlations in the opposite direction. Notably, the Scz ∩ CanUD ∩ Smk and Scz ∩ CanUD | Smk subsets were negatively genetically correlated with executive function, while the Scz ∩ Smk | CanUD subset was positively correlated and the CanUD ∩ Smk | Scz subset was not significantly correlated, suggesting a pivotal role of the intersection of CanUD and Scz, regardless of Smk, on executive functioning.

We also created polygenic scores (PGS) from each SNP subset in the European ancestry data and tested their associations with a range of health-related phenotypes in the BioVU biobank. In line with the genetic correlations in GNOVA, the PGS for the convergent subset of SNPS (Scz ∩ CanUD ∩ Smk) showed the strongest associations overall with most subsets of traits (**Figure 4A**), especially suicide attempt, psychosis, PTSD, conduct disorders, antisocial/borderline personality disorder, bipolar disorder, and alcohol-related disorders, among other psychiatric phenotypes (**Figure 4B**). Exceptions to this pattern included metabolic and endocrine phenotypes (**Figure 4B**), for which the PGS for the CanUD ∩ Smk | Scz subset had the greatest magnitude of associations with many of these traits, including acidosis, adult failure to thrive, type 2 diabetes, and hyperkalemia.

**Figure 4.**
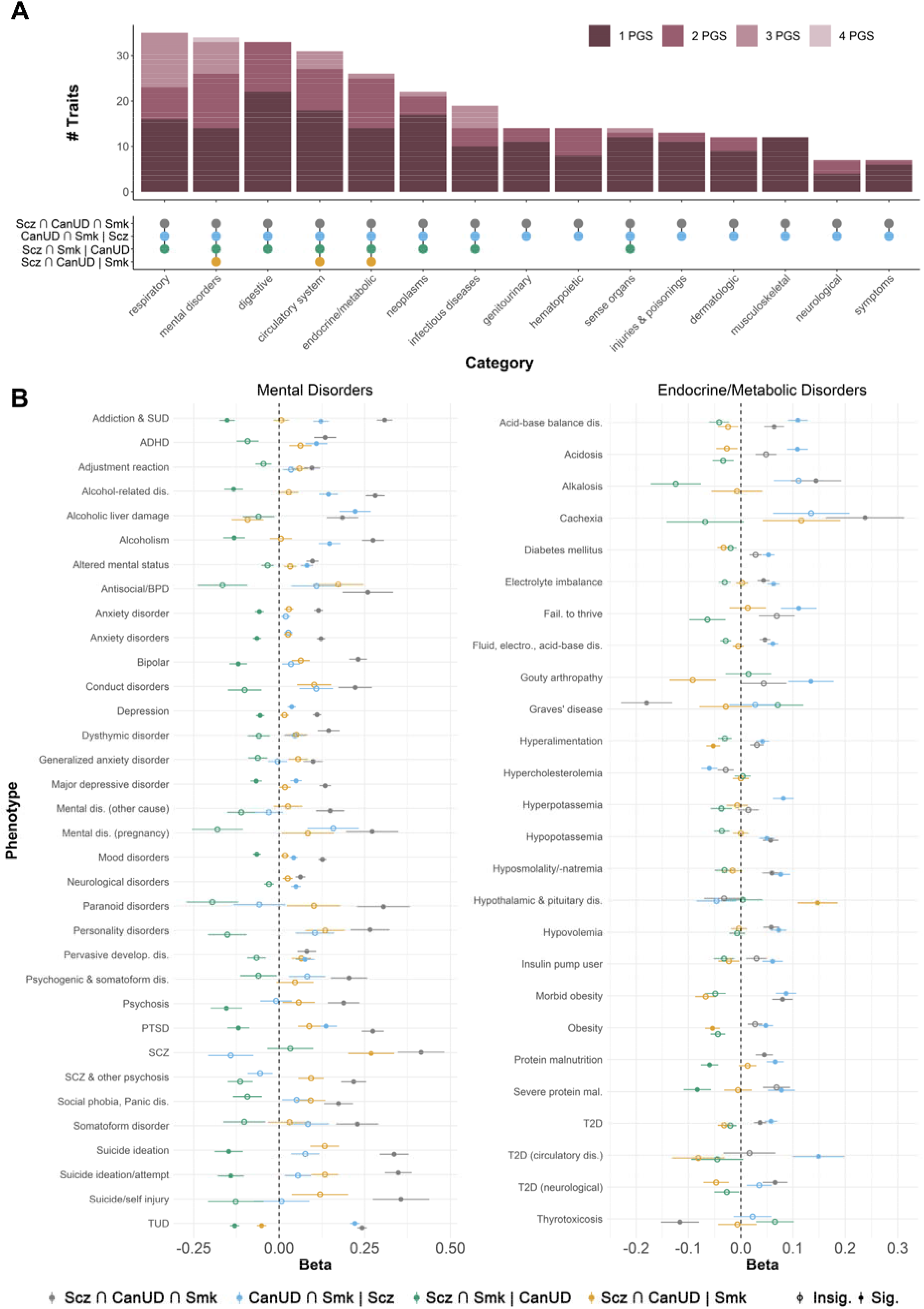
Associations between polygenic scores for SNP subsets from ASSET and health-related phenotypes in the BioVU biobank. Panel A: Upset plot showing the number of phenotypes within different categories associated with one or more PGS. Panel B: Forest plots showing associations between the four ASSET SNP subset PGSs and mental disorders (left panel) and endocrine/metabolic traits (right panel) in BioVU.

### Partitioned genetic covariance analysis

When stratified by broad tissue type, the genetic covariance between CanUD and Scz was significantly enriched for brain tissues in the European ancestry data (*ρ* = 0.029, p = 8.3e-4), while the genetic covariance between Smk and Scz was not significantly enriched for any tissue category (**Figure S1, Table S12**).

## Discussion

The nature of the relationship between cannabis use and schizophrenia is a compelling and fiercely debated question in psychiatry, one that is complicated by the possibility of shared genetic factors and the frequent co-occurrence with tobacco smoking. There are major public health implications associated with a causal effect of cannabis use on schizophrenia risk, so a resolution of this question is important. Here, we describe the largest genome-wide, cross-ancestry and cross-disorder analyses of cannabis use disorder (CanUD), tobacco smoking (Smk), and schizophrenia (Scz) to date.

Our analyses revealed three key findings. First, CanUD and Smk are both genetically correlated with Scz, and this was consistent in both the European and African ancestry datasets. However, CanUD and Scz showed a greater degree of genetic overlap than Smk and Scz. Second, causal inference analyses suggested evidence of bidirectional causality for genetic liability to Scz, CanUD, and Smk, albeit in the presence of horizontal pleiotropy. Third, genomic loci that comprise the intersection between CanUD and Scz are associated with other mental health conditions and executive functioning.

In causal inference analyses that accounted for both correlated and uncorrelated forms of horizontal pleiotropy, we saw evidence for bidirectional causal relationships between all three phenotypes. We found evidence of horizontal pleiotropy for all trait pairs through the MR-PRESSO global test, but again found significant bidirectional causal estimates even after removing outlier SNPs for horizontal pleiotropy. Collectively, these results support causal links between CanUD, Smk, and Scz, although it is worth noting that the MR-Egger test did not support any causal relationships except for genetic liability for Smk causing CanUD. Convergent evidence from additional sources (especially longitudinal, prospective cohort studies) are needed^53^, especially in light of conflicting results from epidemiological studies^5,6,54^ and the limitations (and assumptions) associated with genetic methods of causal inference^55^.

Over 200 loci had convergent genome-wide significant effects on CanUD, Smk and Scz. The strongest convergent locus was on chromosome 8, with the lead SNP being a brain eQTL for *EPHX2, CHRNA2*, and *CCDC25*. While *CHRNA2*, a nicotinic cholinergic receptor (nAChR), seems an intuitive finding for Smk, this locus was most strongly associated with Scz and CanUD (**Figure 2**), and the top lead variant in all recent CanUD GWASs has mapped to this locus^12,17,42^. The role of cholinergic disturbance in positive^56^ symptoms and cognitive symptoms ^57^ of Scz raise the potential for use of nChR agonists for treatment of comorbid Scz, CanUD and Smk^58^. *EPHX2* encodes soluble epoxide hydrolase (sEH), the overexpression of which has been implicated in Scz^59^ and other diseases with a neuroinflammatory component (e.g., Alzheimer’s Disease). There is evidence for synergy between sEH and fatty acid amide hydrolase (FAAH^60^), which metabolizes endogenous cannabinoids and the inhibition of which is being evaluated for the treatment of pain. Given the emerging and paradoxical role of CanUD and a proinflammatory state^61^, the role of *EPHX2* at the intersection of these disorders is intriguing.

Variants previously implicated in metabolic phenotypes emerged from the Scz ∩ CanUD| Smk subset. For instance, the lead SNP rs9924686 in *TAOK2* was negatively associated with CanUD and Scz but not Smk and has been implicated in numerous prior GWAS of metabolic traits^62,63^. Further, while PGS derived from this subset were associated with both psychiatric and metabolic traits, the direction of association differed between this subset and the polygenic score of the fully convergent subset for metabolic but not psychiatric phenotypes (**Figure 4**).

Genetic predisposition for executive functioning was negatively correlated with the subset of fully convergent variants (Scz ∩ CanUD ∩ Smk), as well as those in the Scz ∩ CanUD | Smk subset, but positively associated with the other two subsets (i.e., where effects diverged for either CanUD or Scz), suggesting that only variation shared by CanUD *and* Scz related to lower executive functioning. Executive functioning deficits are a defining feature of Scz^52^ and a broad range of substance use disorders^51,64,65^, consistent with our findings. Executive functioning deficits have also been implicated in a broader range of mental health conditions^51^, which also aligns with our observation that variants influencing CanUD and ScZ, regardless of their effects of Smk, appear to index serious psychiatric comorbidity. Notably, while executive functioning is related to educational attainment^51^, the pattern of associations between convergent subsets of variants and educational attainment appeared to be quite different – for instance, any subset where CanUD and Smk diverged was associated with greater educational attainment, suggesting that educational attainment was more closely related to convergent signals for CanUD and Smk, rather than Scz. Thus, our study implicates the genetic liability to lower executive functioning as a common mechanism undergirding CanUD and Scz.

The genetic correlation between CanUD and Scz (r_g_ = 0.37, SE = 0.02) was significantly greater (p_diff_ = 6.5e-18) than that between Smk and Scz (r_g_ = 0.17, SE = 0.02) in the European ancestry data (with a similar but non-significant pattern in the African ancestry data: r_g_(CanUD, Scz) = 0.61, SE = 0.14 vs. r_g_(Smk, Scz) = 0.34, SE = 0.15). This suggests a greater proportion of shared genetic effects for CanUD and Scz than for Smk and Scz. When we partitioned the genetic covariance between phenotype pairs (Scz and CanUD, and Scz and Smk) into broad tissue types, the genetic covariance of CanUD and Scz was enriched for brain tissue, while the genetic covariance between Smk and Scz was not significantly enriched in any tissue category. These results are consistent with an overall pattern of findings in our study: the degree of genetic overlap, and the extent to which it is enriched in meaningful biological categories, is greater for CanUD and Scz than for Smk and Scz.

Our analyses of African ancestry data increase the generalizability of our findings. However, the smaller sample size of the individual African ancestry GWASs and limited available data for follow-up analyses (e.g., annotation files for partitioned genetic covariance analyses) constrained the extent to which we were able to accomplish our goal of equitable analyses. The genetic correlation between CanUD and Scz was substantially larger in the African ancestry data (r_g_ = 0.610, SE = 0.140, p = 1.41e-5) than in the European ancestry data (r_g_ = 0.373, SE = 0.023, p = 2.97e-60), albeit with a much larger standard error, suggesting that with increasing sample size, there could be considerable opportunity to identify pleiotropic loci.

Several other limitations applied to our study. First, while early age of cannabis initiation and use of high-potency cannabis have been suggested as risk factors for Scz, we did not have data available on potency and did not include age at first use in our analyses, as the only available GWAS for this phenotype was relatively underpowered and had non-significant SNP-heritability^66^. Similarly, we were unaware of any GWAS of cannabis consumption (i.e., heaviness or frequency of use). Another limitation is that the individual GWAS likely contain comorbid cases (e.g., a SCZ case with co-occurring CanUD), and this could artificially inflate our estimates of genetic correlations. Furthermore, cross-trait assortative mating has been shown to bias genetic correlations (e.g., between alcohol use disorders and schizophrenia)^67^, although the extent to which this could be affecting estimates of correlation between Scz, CanUD, and Smk specifically has not yet been quantified. Finally, there may be some sample overlap among the different GWAS (especially for CanUD and Smk), which could have inflated our MR results (but not CAUSE, which accounts for sample overlap).

Overall, our results add to the body of literature suggesting that both Smk and CanUD may be important predisposing factors as well as sequela of Scz. We demonstrate that the relationship between Smk, CanUD, and Scz may be due to both correlated genetic and reciprocal causal effects. Further, we identify executive functioning as a potential phenotype that links genetic liability for CanUD and Scz. While cigarette use is generally decreasing^68^, nicotine exposure through vaping is increasing^26,69^ and cannabis legalization and use are becoming more widespread worldwide^70^. As substance use policies and modes of use continue to change, it is important to carefully monitor epidemiologic trends in mental health conditions, especially schizophrenia and other psychotic disorders, and consider targeted interventions that may benefit individuals with heavy cannabis and tobacco use.

## Methods

### Genome-wide summary statistics

We used summary statistics from the largest available GWAS of each trait: Scz, CanUD and tobacco smoking.

- *Schizophrenia (Scz):* We used data from the most recent Psychiatric Genomics Consortium (PGC) Schizophrenia genome-wide association study (GWAS) meta-analysis of individuals of European ancestry (N = 161,405; N_cases_ = 67,390)^32^. We also analyzed summary statistics from a GWAS meta-analysis of schizophrenia in African ancestry individuals (N = 15,846; *N*_cases_ = 7509), from the Cooperative Studies Program (CSP) #572 and the Genomic Psychiatry Cohort^71^.
- *Cannabis use disorder (CanUD):* We used data from Levey et al.’s recent GWAS meta-analysis of cannabis use disorder^17^, which combined data from the Million Veteran Program, the Psychiatric Genomics Consortium, the Lundbeck Foundation Initiative for Integrative Psychiatric Research, and deCODE Genetics (European ancestry N = 886,025; N_cases_ = 42,281; African ancestry N = 120,208; N_cases_ = 19,065).
- *Ever-smoked tobacco regularly (Smk):* We used summary statistics from the GWAS & Sequencing Consortium of Alcohol and Nicotine use (GSCAN) GWAS of self-reported ever/never regular cigarette smoking (European ancestry N = 805,431; N_ever_ = 393,707; African ancestry N = 24,278; N_cases_ = 9,916)^72^. We used the publicly-available set of summary statistics, which does not include data from 23andMe; the sample sizes reported here reflect that exclusion. This phenotype was measured in a variety of ways in different cohorts (e.g., “Have you smoked over 100 cigarettes over the course of your life?”, “Have you ever smoked every day for at least a month?”, “Have you ever smoked regularly?”). We selected this phenotype over others reflecting smoking quantity (cigarettes per day) or dependence because it had the largest sample size and the most genome-wide significant loci of any tobacco-related GWAS and shows considerable overlap with other nicotine use traits; thus, it seems likely that the genetics of Smk would be inclusive of most genetic factors related to tobacco involvement.

We also used genome-wide summary statistics for educational attainment, executive function, risk-taking, and the Townsend Deprivation Index (TDI):

- *Educational attainment:* We used data from a GWAS of educational attainment from Lee et al.^50^ (2018) in individuals of European ancestry (N = 766,345).
- *Executive function:* We used summary statistics from a GWAS of executive function in the European ancestry subset of the UK Biobank by Hatoum et al.^51^ (2023; N = 427,037).
- *Risk-taking:* We used data from a GWAS by Linnér et al.^46^ (2019) of a single item that queried whether someone was a risk-taker. This GWAS was a meta-analysis of the UK Biobank and 10 replication cohorts (N = 466,571).
- *TDI:* We used summary statistics from the Neale Lab GWAS (https://www.nealelab.is/uk-biobank) of the Townsend Deprivation Index (a measure of material deprivation in a region, incorporating data on unemployment, non-car-owning households, non-home-owning households, and household overcrowding) in the European ancestry subset of the UK Biobank (N = 336,798).

### Genome-wide genetic correlation analyses

We used linkage disequilibrium score regression^73,74^ (LDSC) to estimate pairwise genome-wide genetic correlations (r_g_) between Scz, Smk, and CanUD. For the European ancestry summary statistics, we used pre-computed LD scores from the 1000 Genomes Phase 3 European reference panel (available from the LDSC website). For the African ancestry summary statistics, we used pre-computed LD scores from the PanUKBB African ancestry sample (available from https://pan.ukbb.broadinstitute.org/downloads).

We further tested whether genetic correlations were significantly different from each other using a block-jackknife method^74,75^. The block-jackknife method is a resampling approach, where the difference between resampling genetic correlations is used to calculate a jackknife standard error. From this standard error a Z-statistic is estimated and used in a two-tailed Z-test to determine if the difference between two genetic correlations is significantly different from zero (i.e., H_0_: r_g_(Scz, Smk) - r_g_(Scz, CanUD) = 0).

### Causal inference analyses

We tested for causal relationships between Scz, CanUD, and Smk using CAUSE^35^. Compared to traditional Mendelian Randomization methods, CAUSE has the advantage of accounting for correlated horizontal pleiotropic effects (i.e., a genetic instrument is associated with a confounder which is related to both the exposure and the outcome) as well as uncorrelated horizontal pleiotropy. CAUSE uses a less stringent p-value threshold (p < 1e™3) to incorporate data from more variants across the genome. CAUSE constructs two nested models: a sharing model and causal model. Both models allow for horizontal pleiotropic effects; however, only the causal model includes a causal effect parameter (gamma). CAUSE compares the sharing and causal models to each other, to determine which model best fits the data, by estimating the difference in the expected log pointwise posterior density (ΔELPD). CAUSE then computes a z-score from the ΔELPD that can be compared to a normal distribution to obtain a one-sided p-value, which corresponds to a test of the null hypothesis that the sharing model fits the data at least as well as the causal model. Significant p-values therefore indicate the presence of a causal effect, after accounting for pleiotropy.

We performed additional causal inference analyses, including Mendelian Randomization Pleiotropy RESidual Sum and Outlier (MR-PRESSO^36^) analyses to test for horizontal pleiotropy and causal relationships among Scz, CanUD, and Smk. We performed these analyses using the TwoSampleMR R package^76,77^. We required SNP instruments to have p-value ≤ 5e-8 and performed LD-based clumping. We report the results from the MR-PRESSO global test for horizontal pleiotropy, MR-PRESSO test for causality after removing outliers for horizontal pleiotropy, MR-Egger, weighted median, inverse variance weighted, simple mode, and weighted mode tests for causality, heterogeneity tests for the inverse variance weighted and MR-Egger tests, and the MR-Egger pleiotropy test (**Table S3**).

We only performed these analyses using the European ancestry summary statistics, because the African ancestry summary statistics were relatively under-powered for a causal inference analysis, particularly the Scz summary statistics.

### Cross-disorder genome-wide association study meta-analysis

We used ‘Association analysis based on SubSETs’ (ASSET^39^) to combine the GWAS summary data for CanUD, Smk and Scz (separately by ancestry), using the two-tailed meta-analysis approach to obtain a single cross-disorder association statistic. Unlike traditional meta-analysis approaches, ASSET takes into account SNPs with significant effects on multiple disorders even if the effects on the traits are in opposite directions. We used the LDSC genetic covariance intercept to approximate the degree of sample overlap amongst the studies and included it in the ASSET covariance matrix. Default parameters were applied using the ‘h.traits’ function.

We then separated the ASSET results into subsets. Following Lam et al.^40^, we use the following notation for each subset: ∩ represents variant subsets with the same directions of effect (+ or -), and | represents variant subsets whose effect sizes are in the opposite direction of those for one versus the other two traits. We defined four subsets: (1) Scz ∩ CanUD ∩ Smk (i.e., a subset with convergent effects across all 3 traits); (2) Scz ∩ CanUD | Smk (i.e., a subset of variants with convergent effects for Scz and CanUD, but divergent effects for Smk); (3) Scz ∩ Smk | CanUD; and (4) CanUD ∩ Smk | Scz.

For each subset, we used FUMA v1.6.1^78^ for annotation and identification of genome-wide significant risk loci and independent lead SNPs. We used the matching ancestry subset of the 1000 Genomes Project Phase 3^79^ reference panel for clumping and annotation of SNPs (e.g., the African ancestry reference panel for our African ancestry cross-disorder summary statistics). We used the default parameters for FUMA, with “independent SNPs” defined as those with p < 5e-8 and independent of each other with LD r^2^ ≤ 0.6, and “lead SNPs” as independent SNPs which are strictly independent at a more stringent LD r^2^ ≤ 0.1. Genomic risk loci (defined by LD blocks of independent SNPs) that were 250 kb or closer were merged into a single locus.

To perform a cross-ancestry meta-analysis, we used the ancestry-specific one-sided meta-analysis results from ASSET. Unlike the two-tailed approach described above, the one-sided meta-analysis in ASSET is more akin to a traditional meta-analysis and results in one effect size per SNP, regardless of whether the SNP shows divergent directions of effect across traits. We combined the ancestry-specific ASSET results using a sample-size weighted meta-analysis scheme in METAL^80^. As before, we uploaded METAL results to FUMA for clumping and annotation, using the 1000 Genomes Project Phase 3^79^ all ancestries reference panel.

### Identification of novel loci

To determine whether the ASSET meta-analysis revealed any novel loci in the European ancestry data that were not genome-wide significant in the original GWAS (CanUD, Smk, Scz), we used the LDLink package^81^ in R to identify all LD proxy SNPs (r^2^>0.6) for each of the 439 lead pleiotropic SNPs. We then merged these results with the summary statistics for the original CanUD, Smk, and Scz GWASs to determine whether the locus had been identified as genome-wide significant in any of the original GWASs.

### Genetic correlations with other relevant phenotypes

After defining SNP subsets using ASSET, we used GeNetic cOVariance Analyzer (GNOVA^49^) to estimate genetic covariances (ρ_g_) and correlations (r_g_) between the SNP subsets and relevant phenotypes in the European ancestry data. For all subsets, the effect estimate was aligned with the direction of effect for CanUD, for ease of interpretation. It was unclear how best to weight the estimate for each subset; following the example of Lam et al.^40^, we used the largest absolute effect size from the three phenotypes as SNP weights in each subset (flipping the sign of the estimate as necessary, to align with the direction of effect for CanUD).

### Polygenic scores of ASSET-derived SNP subsets and associations in BioVU

We created polygenic scores for each ASSET-derived SNP subset in the European ancestry subset of the BioVU biobank (N=72,225)^82,83^. We fitted a logistical regression model to each of 1,338 case/control phenotypes (“phecodes”) to estimate the odds of diagnosis given each PGS. Models were adjusted for sex, median age of the longitudinal electronic health records, and the first 10 PCs. Analyses were conducted using the PheWAS version 0.99.5-2 R package. Phecodes were excluded from the analysis if they did not have at least two International Disease Classification codes mapping to a PheWAS disease category (Phecode Map 1.2; https://phewascatalog.org/phecodes) and had less than 100 cases. The disease phenotypes included 145 circulatory system, 123 genitourinary, 118 endocrine/metabolic, 125 digestive, 118 neoplasms, 91 musculoskeletal, 85 sense organs, 73 injuries & poisonings, 68 dermatological, 76 respiratory, 69 neurological, 64 mental disorders, 42 infectious diseases, 42 hematopoietic, 34 congenital anomalies, 34 symptoms, and 31 pregnancy complications. The phenome-wide significance threshold was set at a Bonferroni-adjusted threshold of *p* ≤ 3.62e-5.

### Partitioned genetic covariance analyses

We used GNOVA^49^ to partition the genetic covariance (ρ_g_) between CanUD, Smk, and Scz into salient annotation categories. These included tissue-specific functionality (GenoSkyline annotations, which are tissue-specific functional regions defined by integrating high-throughput epigenetic annotations). GNOVA is robust to potential sample overlap between summary statistics. We applied Bonferroni correction for multiple testing across all 3 trait pairs (CanUD ∼ Smk, CanUD ∼ Scz, and Smk ∼ Scz) and 7 tissue types tested (e.g., we corrected for 3 x 7 = 21 tests, for an α = 0.002.) We only performed these analyses using the European ancestry summary stats, as the annotation data was derived using European ancestry samples.

## Data Availability

All data produced in the present study will be made available upon publication at the following link: https://github.com/WashU-BG/CanUD_Smk_Scz

