## Supplemental Figure 1 for "Cross-ancestry genetic investigation of schizophrenia, cannabis use disorder, and tobacco smoking"

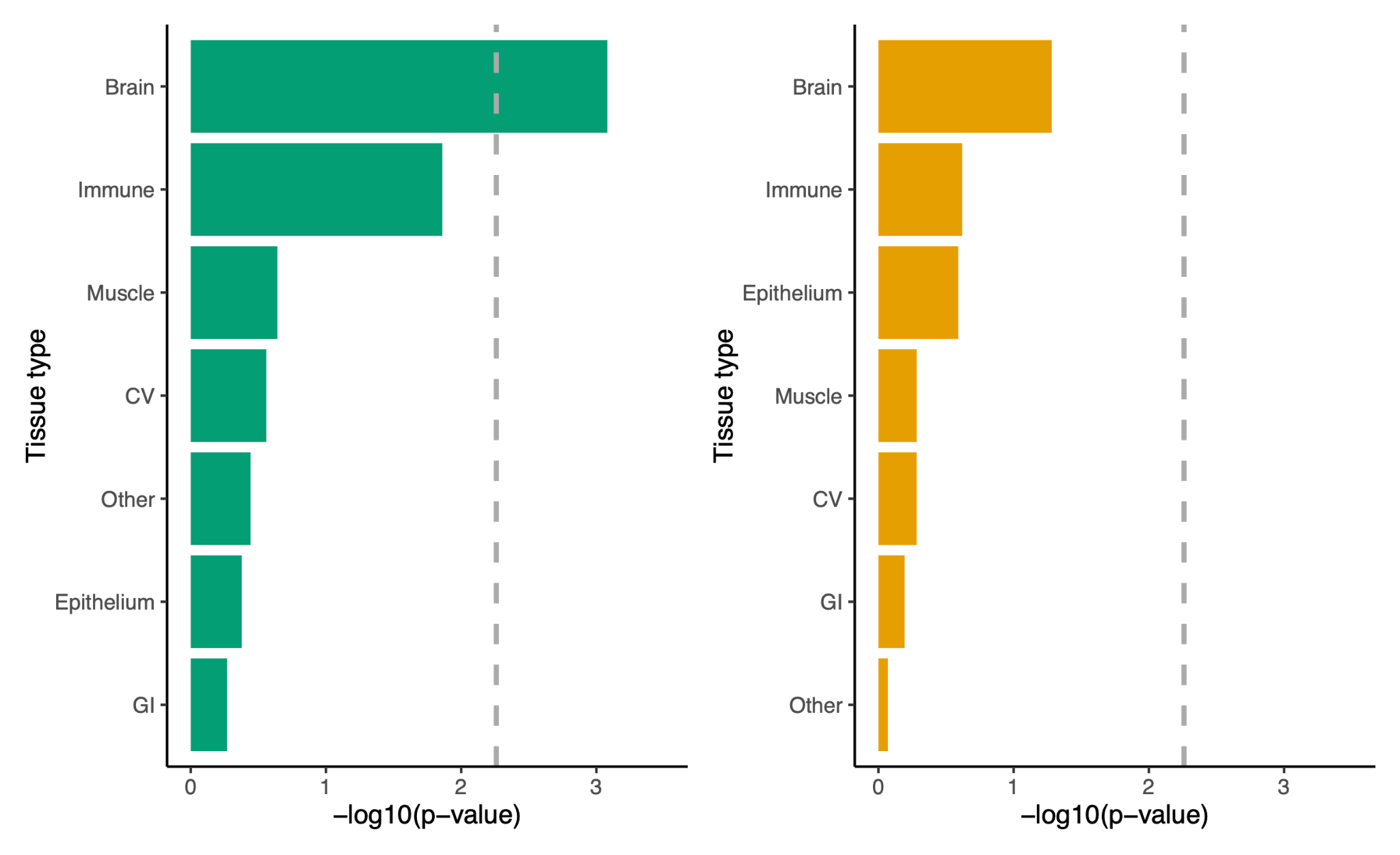

**Supplemental Figure 1. Enrichment of the genetic covariance between cannabis use disorder and schizophrenia (green) and tobacco smoking and schizophrenia (orange) in broad tissue types.** Statistical significance after Bonferroni correction for multiple testing is indicated by the dashed gray line.
